# Lymphocyte count is a universal predictor to the health status and outcomes of patients with coronavirus disease 2019 (COVID-19): A systematic review and meta-regression analysis

**DOI:** 10.1101/2021.08.02.21261505

**Authors:** Kuan-Lang Lai, Fu-Chang Hu, Fang-Yu Wen, Ju-Ju Chen

## Abstract

**Background:** This study aimed to evaluate the prediction capabilities of clinical laboratory biomarkers to the prognosis of COVID-19 patients.

**Methods:** Observational studies reporting at least 30 cases of COVID-19 describing disease severity or mortality were included. Meta-data of demographics, clinical symptoms, vital signs, comorbidities, and 14 clinical laboratory biomarkers on initial hospital presentation were extracted. Taking the outcome group as the analysis unit, meta-regression analysis with the generalized estimating equations (GEE) method for clustered data was performed sequentially. The unadjusted effect of each potential predictor of the three binary outcome variables (i.e., severe vs. non-severe, critically severe vs. non-critically severe, and dead vs. alive) was examined one by one by fitting three series of simple GEE logistic regression models due to missing data. The worst one was dropped one at a time. Then, a final multiple GEE logistic regression model for each of the three outcome variables was obtained.

**Findings:** Meta-data was extracted from 76 articles, reporting a total of 26,627 cases of COVID-19. Patients were recruited across 16 countries. The number of studies (patients) included in the final models of the analysis for severity, critical severity, and mortality was 38 studies (9,764 patients), 21 studies (4,792 patients), and 24 studies (14,825 patients), respectively. After adjusting for the effect of age, lymphocyte count mean or median ≤ 1.03 (estimated hazard ratio [HR] = 46.2594, p < 0.0001), smaller lymphocyte count mean or median (HR < 0.0001, p = 0.0028), and lymphocyte count mean or median ≤ 0.8714 (HR = 17.3756, p = 0.0079) were the strongest predictor of severity, critical severity, and mortality, respectively.

**Interpretation:** Lymphocyte count should be closely watched for COVID-19 patients in clinical practice.

## Introduction

Although numerous treatment options and vaccines are authorized for COVID-19,^1, 2^ the situation of a global pandemic is still continuing. Each day over four hundred thousand new cases are identified even in time of July 2021.^3^ COVID-19, the illness caused by infection with SARS-nCoV2,^4^ is spreading since December 2019 from Wuhan, China, and has accumulated more than 192 million cases and more than 4 million deaths in over 219 countries, area or territories up to 23 July 2021.^5^ During pandemic period medical care system started to overwhelmed in bunch communities no matter from economically developed or underdeveloped regions.^6–13^ How to use simple tools to differentiate, triage patients is crucial.

Several laboratory data have been identified as predictors for disease severity or mortality of COVID-19 patients, e.g., lymphocyte count, neutrophil/lymphocyte ratio (NLR), lactate dehydrogenase (LDH), D-dimer, C-reactive protein (CRP), procalcitonin (PCT).^14–26^ However, since many studies were conducted at the same region during a short period of time,^14–22, 26, 27^ potential bias of subject duplication cannot be ruled out for the following meta-analysis.^28–34^ Additionally the relative strength of broader spectrum lab data for their prediction capability has not been explored on a head-to-head basis. This study aimed to investigate whether laboratory data at hospital presentation play a role in distinguishing severity or predicting mortality for COVID-19 patients and to explore the relative significance of these predictors across regions.

## Research in context

### Evidence before This Study

Plentiful tools, e.g., demographics, symptoms, vital signs, comorbidity, imaging, and lab data have been explored to their prediction capability for COVID-19 patients. Several lab data such as lymphocyte, NLR, LDH, CRP, PCT have been identified to distinguish the severity or to predict the survival of COVID-19 patients. However, most of the meta-analysis research conducted in a specific region at the early stage of the pandemics and subject duplication concerns cannot be ignored due to the large amount papers published within a short period. In addition, the relative strength of these lab items for predictions has not been tested under a broader spectrum which including severity, critical severity, and mortality and across different regions.

### Added Value of This Study

Our study proves that several lab data (e.g., WBC count, lymphocyte count, neutrophil count, platelet count, LDH, D-dimer, CRP, and PCT) of COVID-19 patients at initial hospital presentation holds the values to differentiate disease severity (severe vs. non-severe, critically severe vs. non-critically severe) and to predict the final consequence, mortality (dead vs. alive). After comparisons lymphocyte count was the most powerful predictor among the fourteen explored lab data.

### The implication of All Available Evidence

Early observing essential lab data for a COVID-19 patient can help to scan the health situation, triage the patient, aid for clinical judgment, predict the severity of disease, and from a pragmatic aspect to allocate medical resources appropriately. Under pandemic condition whereas medical resource is constrained, routine lab testing, which is relatively easy access, self-explanation, cost-effectiveness, could be a valuable tool to help for compacting disease. How to maintain or improve good immunity levels for the general population in daily life can be a crucial strategy to stakeholders in facing life-threatening infectious diseases such as COVID-19.

## Methods

### Literature Search Strategy

We used the search terms “COVID-19”, “2019-nCoV”, and “coronavirus” in the search field “Title/Abstract”, at the electronic databases: MEDLINE, and EMBASE. The searches were completed on 10 October 2020.

### Inclusion and Exclusion Criteria

The eligibility criteria for the inclusion of literature in the meta-analysis were as follows: (1) the literature is the original research; (2) the literature was a study with COVID-19 confirmed patients; (3) the literature was published in English with the full text available; (4) source of subjects, recruitment situation were clearly stated. Literature was excluded from the meta-analysis when (1) disease severity or survival status was not well defined; (2) pediatric study or particular subject group, e.g., specific disease apart from COVID-19; (3) desired lab data at hospital administration for COVID-19 was not available which fourteen lab items were: white blood cell (WBC), neutrophil, lymphocyte, neutrophil to lymphocyte ratio (NLR), platelet, alanine aminotransferase (ALT), aspartate aminotransferase (AST), creatinine, D-dimer, C-reactive protein (CRP), procalcitonin (PCT), lactate dehydrogenase (LDH), and hypersensitive troponin I (hs-cTnI); (4) subjects number below thirty; (5) research subjects may duplicate from other studies after investigation of the sites and the recruitment period. Under condition (5), the study with utmost information by calculating (the number of study subjects) × (number of lab data items)] was selected.

At the initial stage, after duplicates were removed, 1,126 records were identified from MEDLINE or EMBASE databases. Of the leaving records, after the title and abstract review, 660 documents were excluded. The leaving 466 articles were carefully and detailed evaluated. At last, 390 articles were excluded, because the studies did not meet the criteria we have set. Finally, a total of 76 studies with 26,627 patients were included in qualitative synthesis (Figure 1). Among the 76 studies, based on the features of data, a total of 38 studies, 21 studies, and 24 studies were incorporated in the analysis for severity, critical severity, and mortality respectively. After all, a total of 35 studies, 15 studies, and 19 studies were presented in the meta-regression analyses, respectively.

**Figure 1:**
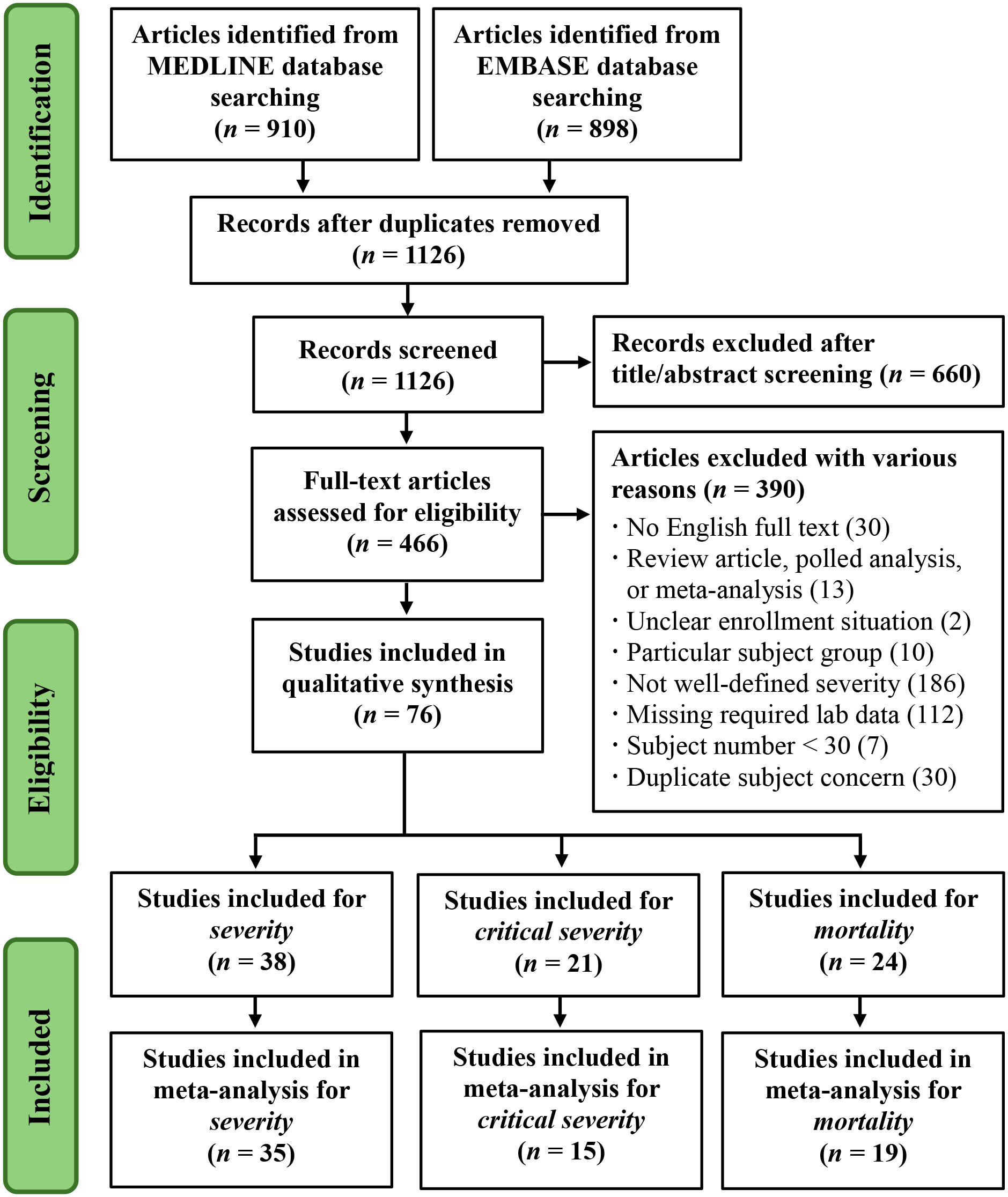
Flow diagram of literature search and study selection (PRISMA chart).

### Data Extraction

The following data were extracted from the qualified studies: first author, year/month of the publication, location (city-country), hospital name, definition of disease severity, subject number, number of COVID-19 patients in each health status, age, male to female ratio, vital sign, clinical feature (12 symptoms), comorbidity (any; 8 main diseases), and desired 14 lab data [Appendix I, Appendix II). Lab data on the initial hospital presentation were classified as blood routine, blood biochemistry, coagulation functions, inflammatory markers, myocardial injury markers (see Appendix II).

The primary outcome measures were to compare the level of laboratory data and their impact on different health outcomes (non-severe vs. severe, non-critically severe vs. critically severe, and alive vs. dead) after adjusting the effects of other covariates.

### Statistical Analysis

Statistical analysis was performed using the R 4.1.0 software (R Foundation for Statistical Computing, Vienna, Austria). Two-sided *p* value ≤ 0.05 was considered statistically significant. We chose the outcome groups in the collected studies as the analysis unit ― instead of the collected studies themselves ― in this meta-analytical study. The distributional properties of continuous variables were expressed by mean ± standard deviation (SD), median, interquartile range (IQR), and categorical variables were presented by frequency and percentage (%). In univariate analysis, the unadjusted effect of each potential risk factor, prognostic factor, or predictor of the three binary outcome variables (i.e., severe vs. non-severe, critically severe vs. non-critically severe, and dead vs. alive) was examined respectively using the Wilcoxon rank-sum test, Chi-square test, and Fisher’s exact test as appropriate for the data type. Next, multivariate analysis was conducted by fitting the logistic regression models to estimate the adjusted effects of potential risk factors, prognostic factors, or predictors on the three binary outcome variables (i.e., severe vs. non-severe, critically severe vs. non-critically severe, and dead vs. alive) respectively with the generalized estimating equations (GEE) method. The GEE method was used to account for the correlation between the two outcome groups within a collected study.^35^ Computationally, we used the geeglm function (with the specified “exchangeable” correlation structure and the default robust estimator of standard error) of the geepack package^36, 37^ to fit GEE logistic regression models for the three sets of correlated binary responses (i.e., severe vs. non-severe, critically severe vs. non-critically severe, and dead vs. alive) respectively in R.

To ensure a good quality of analysis, the model-fitting techniques for (1) variable selection, (2) goodness-of-fit (GOF) assessment, and (3) regression diagnostics and remedies were used in our GEE logistic regression analyses. All the univariate significant and non-significant relevant covariates (listed in Appendix II) were put on the variable list to be selected. However, each of the collected studies selectively reported the potential risk factor, prognostic factor, or predictor of the three binary outcome variables. If we wanted to assess simultaneously the effects of all the relevant covariates (listed in Appendix II), then the number of studies without missing values would be very few. Thus, our meta-regression analysis was performed by fitting a series of simple GEE logistic regression models and then dropping the worst one at a time to maximally use all the available information. Then, a final multiple GEE logistic regression model for each of the three outcome variables was obtained. Any discrepancy between the results of univariate analysis and multivariate analysis was likely due to the variation in the number of studies without missing values or the confounding effects of uncontrolled covariates in univariate analysis.

The GOF measures, including the estimated area under the receiver operating characteristic (ROC) curve (also called the *c* statistic) and adjusted generalized *R*^2^, and the Hosmer-Lemeshow GOF test were examined to assess the GOF of the fitted GEE logistic regression model. The value of the *c* statistic (0 ≤ *c* ≤ 1) ≥ 0.7 suggests an acceptable level of discrimination power. Larger *p* values of the Hosmer-Lemeshow GOF test imply better fits of logistic regression model.

Simple and multiple generalized additive models (GAMs) were fitted to draw the GAM plots for detecting nonlinear effects of continuous covariates and then for identifying the appropriate cut-off point(s) to discretize continuous covariates, if necessary, during the above variable selection procedure. Computationally, we used the vgam function of the VGAM package with the default values of the smoothing parameters (e.g., s(age, df=4, spar=0) for the cubic smoothing splines) to fit the GAMs for our binary responses, and then used the plotvgam function of the same package to draw the GAM plots for visualizing the linear or nonlinear effects of continuous covariates in R.^36, 38, 39^ If a separation or high discrimination problem occurred in logistic regression analysis, we fitted the Firth’s bias-reduced logistic regression model using the logistf function of the logistf package in R.^40^ Finally, the statistical tools of regression diagnostics for residual analysis, detection of influential cases, and check of multicollinearity were applied to discover any model or data problems. The values of the variance inflating factor (VIF) ≥ 10 in continuous covariates or VIF ≥ 2.5 in categorical covariates indicate the occurrence of the multicollinearity problem among some of the covariates in the fitted logistic regression model.

## Results

### Literature Search and Study Characteristics

Based on the search strategy, 76 articles were included in the qualitative synthesis^10, 14, 16–25, 41–104^ including 26,627 COVID-19 confirmed patients (Figure 1, Appendix I). The number of studies (patients) included for the analysis of severity, critical severity, and mortality was 38 studies (9,764 patients), 21 studies (4,792 patients), and 24 studies (14,825 patients) respectively (Figure 1, Appendix I). Patient demographics, clinical features, comorbidities, lab items for each health status were shown (Appendix, Appendix II). All of the selected articles were published in 2020 with patient sizes ranged from 38 to 4,035 subjects. All of the articles were retrospective, observational studies with COVID-19 patients recruited between 1 December 2019 and 27 Jun 2020, from sixteen countries. The majority of studies were conducted in China (46 studies, 60.5%), followed by Italy (5 studies, 6.6%) and the US (5 studies, 6.6%). Most of the subjects come from China (13,483 patients, 50.6%), followed by Spain (4,035 patients, 15.2%) and US (2,691 patients, 10.1%). Except for 21 sole mortality studies, 55 studies incorporated disease severity definitions (Appendix I). Most (49, 89.1%) of the studies were based on the WHO interim guidance^105^ or national guidance modified from the WHO principles^91, 106, 107^, followed by the American Thoracic Society Guideline (5, 9.1%) and the International Guideline for Community-Acquired Pneumonia (1, 1.8%). Since the contents of the above guidelines were similar, they were all included in the analysis. In general, disease severity is classified into four types: mild, moderate, severe, and critically severe. Severe: Meet any of the following (1) Shortness of breath, RR>30 times per minute; At room air, SpO2 lower than 93%; (3) The partial pressure of Arterial blood oxygen (PaO2)/the fraction of inspired oxygen (FiO2) ≤ 300mmHg; (4) CT chest imaging shows that lung damage develops significantly within 24 to 48 hours. Critically severe: Meet any of the following (1) Respiratory failure requiring mechanical ventilation; (2) Signs of septic shock; Multiple organ failure requiring ICU admission. For comparison purposes in this study, the subjects in the mild and the moderate conditions were assembled into the non-severe group; subjects in the mild, moderate, and severe were assembled into the non-critical group. There were therefore six health outcomes classified into three pairs in this study: severe vs. non-severe, critically severe vs. non-critically severe, dead vs. alive.

### Patient Demographics, Clinical Characteristics, Comorbidities, and Lab Data

Summary statistics of the demographics, clinical characteristics, comorbidities, and laboratory data of the COVID-19 patients on initial hospital presentations for the assessment of severity, critical severity, and mortality were shown in Appendix II-1, Appendix II-2, and Appendix II-3, respectively.

Not every desired lab parameter was collected for each study ― for example, NLR and hs-cTnI were rarely reported. Most of the lab data had statistical significance (all p < 0.05) between the two groups except less collected parameters to the disease severity (Appendix II-1, Appendix II-2); ALT, total bilirubin to the mortality (Appendix II-3).

### Predictors for Severity (Severe vs. Non-severe)

Results of univariate analyses of the predictors for severity (severe vs. non-severe) were shown in Table 1-A. As less effect lab items judged by the AUC of ROC kicked out from the model, more and more arms, from *m* = 14 to *m* = 70 at the different runs, were recruited in the meta-regression analysis. At the final run (seventh, *m* = 70) only lymphocyte count (AUC = 0.938) or lymphocyte count ≤ 1.03 or > 2.06 (0.929) and age (AUC = 0.855) or age > 55.02 (AUC = 0.800) existed in the final univariate analyses. Table 1-B listed the result of multivariate analysis for the prediction of severity (severe vs. non-severe). Age mean or median > 55.02 presented a higher risk to the severity (estimated hazard ratio [HR] = 5.7921, *p* = 0.0058) while lymphocyte count mean or median ≤ 1.03 or > 2.06 showed a strong risk to the severity (HR = 46.2594, *p* < 0.0001).

**Table 1-A:** Univariate analyses of the predictors for severity (severe vs. non-severe) by fitting a series of simple logistic regression models with the generalized estimating equations (GEE) method (assuming an ‘exchangeable’ working correlation structure).

| Order<sup>1</sup> | Covariate | AUC of ROC | 95% C.I. of<br>AUC of ROC | Residual<br>Deviance | R<sup>2</sup> |
| --- | --- | --- | --- | --- | --- |
| **Run 1: $m = 10$** |  |  |  |  |  |
| 1 | CRP mean or median | 1.000 | 1–1 | 0.0000 | 1.0000 |
| 2 | D-dimer mean or median | 1.000 | 1–1 | 0.0000 | 1.0000 |
| 3 | Lymphocyte count mean or median | 1.000 | 1–1 | 0.0000 | 1.0000 |
| 4 | Neutrophil count mean or median $\leq 2.50$ or $> 3.74$ | 1.000 | 1–1 | 0.0000 | 1.0000 |
| 5 | Platelet count mean or median $\leq 194.99$ or $> 232.94$ | 1.000 | 1–1 | 0.0000 | 1.0000 |
| 6 | Age mean or median | 0.920 | 0.736–1 | 6.8134 | 0.6745 |
| 7 | Lactate dehydrogenase mean or median $> 214.99$ | 0.900 | 0.704–1 | 5.4067 | 0.7609 |
| 8 | PCT mean or median $> 0.05$ | 0.900 | 0.704–1 | 5.4067 | 0.7609 |
| 9 | White blood cell count mean or median $\leq 4.21$ or $> 5.72$ | 0.900 | 0.704–1 | 5.4067 | 0.7609 |
| 10 | Age mean or median $> 56.50$ | 0.800 | 0.560–1 | 8.3758 | 0.5631 |
| 11 | Percentage of male gender | 0.560 | 0.153–0.967 | 13.7379 | 0.0166 |
| **Run 2: $m = 14$** |  |  |  |  |  |
| 1 | CRP mean or median | 1.000 | 1–1 | 0.0000 | 1.0000 |
| 2 | D-dimer mean or median | 1.000 | 1–1 | 0.0000 | 1.0000 |
| 3 | Lymphocyte count mean or median | 1.000 | 1–1 | 0.0000 | 1.0000 |
| 4 | Neutrophil count mean or median $\leq 2.39$ or $> 3.70$ | 1.000 | 1–1 | 0.0000 | 1.0000 |
| 5 | Age mean or median | 0.939 | 0.821–1 | 8.6161 | 0.7165 |
| 6 | Age mean or median $> 56.05$ | 0.857 | 0.676–1 | –<sup>2</sup> | 0.5670 |
| 7 | Lactate dehydrogenase mean or median > 224.20 | 0.857 | 0.659–1 | 11.4833 | 0.5763 |
| 8 | Platelet count mean or median $\leq 197.80$ or > 236.14 | 0.786 | 0.557–1 | 14.4041 | 0.4007 |
| **Run 3: $m = 16$** |  |  |  |  |  |
| 1 | CRP mean or median | 1.000 | 1–1 | 0.0000 | 1.0000 |
| 2 | Lymphocyte count mean or median | 1.000 | 1–1 | 0.0000 | 1.0000 |
| 3 | D-dimer mean or median | 0.984 | 0.941–1 | 4.3995 | 0.8945 |
| 4 | Age mean or median | 0.953 | 0.862–1 | 8.8730 | 0.7529 |
| 5 | Lactate dehydrogenase mean or median | 0.953 | 0.851–1 | 10.2592 | 0.7004 |
| 6 | Age mean or median > 55.64 | 0.875 | 0.715–1 | 10.0080 | 0.7103 |
| 7 | Neutrophil count mean or median > 3.71 | 0.875 | 0.702–1 | 12.0566 | 0.6252 |
| **Run 4: $m = 18$** |  |  |  |  |  |
| 1 | CRP mean or median | 1.000 | 1–1 | 0.0000 | 1.0000 |
| 2 | Lymphocyte count mean or median | 1.000 | 1–1 | 0.0000 | 1.0000 |
| 3 | D-dimer mean or median | 0.988 | 0.953–1 | 4.4012 | 0.9077 |
| 4 | Lactate dehydrogenase mean or median | 0.963 | 0.882–1 | 10.5782 | 0.7334 |
| 5 | Age mean or median | 0.938 | 0.834–1 | 10.4480 | 0.7377 |
| 6 | Age mean or median > 55.26 | 0.889 | 0.745–1 | 10.4311 | 0.7383 |
| **Run 5: $m = 28$** |  |  |  |  |  |
| 1 | Lymphocyte count mean or median | 0.929 | 0.820–1 | 19.4244 | 0.6663 |
| 2 | Age mean or median | 0.888 | 0.766–1 | 25.0481 | 0.5179 |
| 3 | CRP mean or median > 27.27 | 0.821 | 0.675–0.968 | 26.1745 | 0.4844 |
| 4 | Age mean or median > 57.33 | 0.750 | 0.584–0.916 | 31.4428 | 0.3087 |
| 5 | D-dimer mean or median > 0.58 | 0.750 | 0.584–0.916 | 31.4428 | 0.3087 |

| Run 6: $m = 56$ | | | | | |
| --- | --- | --- | --- | --- | --- |
| 1 | Lymphocyte count mean or median | 0.952 | 0.897–1 | 30.6650 | 0.7570 |
| 2 | CRP mean or median | 0.926 | 0.851–1 | 47.3394 | 0.5571 |
| 3 | Age mean or median | 0.874 | 0.778–0.970 | 51.0981 | 0.5032 |
| 4 | CRP mean or median > 28.69 | 0.839 | 0.741–0.937 | 49.3143 | 0.5292 |
| 5 | Age mean or median > 56.06 | 0.804 | 0.698–0.909 | 55.4443 | 0.4362 |
| Run 7: $m = 70$ | | | | | |
| 1 | Lymphocyte count mean or median | 0.938 | 0.872–1 | 55.4496 | 0.5973 |
| 2 | Lymphocyte count mean or median $\leq 1.03$ or $> 2.06$ | 0.929 | 0.867–0.990 | 35.8650 | 0.7769 |
| 3 | Age mean or median | 0.855 | 0.762–0.948 | 66.2054 | 0.4750 |
| 4 | Age mean or median > 55.02 | 0.800 | 0.705–0.895 | 69.9269 | 0.4282 |

<sup>1</sup> The simple logistic regression models were listed in the order of the values of the area under the curve (AUC) of Receiver Operating Characteristic (ROC).
<sup>2</sup> The 5 groups with “Age mean or median $\leq 56.06$ years” were all *non-severe* so that the separation or high discrimination problem occurred in fitting the simple logistic regression model with the generalized estimating equations (GEE) method (assuming an ‘exchangeable’ working correlation structure). Then, the `logistf()` function of the `logistf` package was used to fit the Firth’s bias-reduced logistic regression model with the profile penalized log-likelihood method in R. Since the “Residual Deviance” of the Firth’s bias-reduced logistic regression model was not compatible with the others, it was not computed and listed.

**Table 1-B:** Multivariate analysis of the predictors for severity (severe vs. non-severe) by fitting a final multiple logistic regression model with the generalized estimating equations (GEE) method (assuming an ‘exchangeable’ working correlation structure).^1^

| Covariate | Estimated Regression Coefficient | Standard Error | $\chi^2$ Value | *p* Value | Estimated Odds Ratio | 95% Confidence Interval of Odds Ratio |
| --- | --- | --- | --- | --- | --- | --- |
| Intercept | -2.6996 | 0.7028 | 14.7562 | 0.0001 | 0.0672 | 0.0170–0.2666 |
| Lymphocyte count mean or median $\leq 1.03$ or $> 2.06$ | 3.8343 | 0.8442 | 20.6299 | $<0.0001$ | 46.2594 | 8.8435–241.9787 |
| Age mean or median<sup>2</sup> $> 55.02$ | 1.7565 | 0.6364 | 7.6174 | 0.0058 | 5.7921 | 1.6638–20.1634 |

<sup>1</sup> **Goodness-of-fit assessment:** Number of studies, $n = 35$ , number of arms, $m = 70$ (cluster size = 2), the estimated area under the Receiver Operating Characteristic (ROC) curve = $0.968 > 0.7$ with 95% C.I.: 0.928-1 (DeLong).
<sup>2</sup> Although “Age mean or median” beat “Age mean or median $> 55.02$ ” in Run 7 of Table 3-1A (AUC of ROC: 0.855 vs. 0.800), we chose “Age mean or median $> 55.02$ ” in this multiple logistic regression model because the GAM plot of “Age mean or median” indicated that its effect on logit(probability of being severe) flattened off for age’s mean or median $\leq 55.02$ years.

### Predictors for Critical Severity (Critically Severe vs. Non-critically severe)

Results of univariate analyses of the predictors for critical severity (critically severe vs. non-critically severe) were shown in Table 2-A. At the last run (seventh, *m* = 30) only lymphocyte count (AUC = 0.933) and age (AUC = 0.829) or age > 59.82 (AUC = 0.767) were existed in the final univariate analyses. Table 2-A listed the result of multivariate analysis for the predictors to critical severity (critically severe vs. non-critically severe). Age and lymphocyte count were in the final meta-regression model. We found that higher lymphocyte count mean or median had an extremely lower risk of critical severity (HR < 0.0001, *p* = 0.0284) while age mean or median > 59.82 have a higher risk of critical severity (HR = 307.6130, *p* = 0.0009).

**Table 2-A:** Univariate analyses of the predictors for critical severity (critically severe vs. non-critically severe) by fitting a series of simple logistic regression models with the generalized estimating equations (GEE) method (assuming an ‘exchangeable’ working correlation structure).

| Order<sup>1</sup> | Covariate | AUC of ROC | 95% C.I. of AUC of ROC | Residual Deviance | R<sup>2</sup> |
| --- | --- | --- | --- | --- | --- |
| **Run 1: $m = 6$** |  |  |  |  |  |
| 1 | Age mean or median | 1.000 | 1–1 | 0.0000 | 1.0000 |
| 2 | Lactate dehydrogenase mean or median | 1.000 | 1–1 | 0.0000 | 1.0000 |
| 3 | Lymphocyte count mean or median | 1.000 | 1–1 | 0.0000 | 1.0000 |
| 4 | Neutrophil count mean or median | 1.000 | 1–1 | 0.0000 | 1.0000 |
| 5 | CRP mean or median | 0.889 | 0.581–1 | 6.1823 | 0.3993 |
| 6 | D-dimer mean or median | 0.889 | 0.581–1 | 4.2344 | 0.6582 |
| 7 | White blood cell count mean or median | 0.889 | 0.581–1 | 3.7726 | 0.7082 |
| 8 | Platelet count mean or median | 0.667 | 0.013–1 | 8.2068 | 0.0244 |
| **Run 2: $m = 8$** |  |  |  |  |  |
| 1 | Lymphocyte count mean or median | 1.000 | 1–1 | 0.0000 | 1.0000 |
| 2 | Neutrophil count mean or median | 1.000 | 1–1 | 0.0000 | 1.0000 |
| 3 | Age mean or median | 0.938 | 0.764–1 | 4.8792 | 0.7199 |
| 4 | Lactate dehydrogenase mean or median | 0.938 | 0.764–1 | 3.7758 | 0.7990 |
| 5 | White blood cell count mean or median | 0.875 | 0.592–1 | 4.8638 | 0.7211 |
| 6 | CRP mean or median | 0.812 | 0.481–1 | 8.2593 | 0.3974 |
| 7 | D-dimer mean or median | 0.750 | 0.350–1 | 9.8188 | 0.1959 |

| Run 3: $m = 12$ | | | | | |
| --- | --- | --- | --- | --- | --- |
| 1 | Lymphocyte count mean or median | 1.000 | 1–1 | 0.0000 | 1.0000 |
| 2 | Neutrophil count mean or median | 1.000 | 1–1 | 0.0000 | 1.0000 |
| 3 | Age mean or median | 0.944 | 0.816–1 | 5.8139 | 0.7922 |
| 4 | White blood cell count mean or median | 0.917 | 0.738–1 | 6.3748 | 0.7663 |
| 5 | CRP mean or median | 0.833 | 0.595–1 | 11.3907 | 0.4721 |
| 6 | Lactate dehydrogenase mean or median | 0.806 | 0.515–1 | 12.7967 | 0.3650 |
| Run 4: $m = 16$ | | | | | |
| 1 | Age mean or median | 0.938 | 0.807–1 | 10.4603 | 0.6924 |
| 2 | Lymphocyte count mean or median | 0.938 | 0.824–1 | 9.1502 | 0.7428 |
| 3 | Neutrophil count mean or median | 0.938 | 0.807–1 | 10.9031 | 0.6744 |
| 4 | White blood cell count mean or median | 0.844 | 0.624–1 | 13.9161 | 0.5379 |
| 5 | CRP mean or median | 0.781 | 0.543–1 | 17.3436 | 0.3479 |
| Run 5: $m = 24$ | | | | | |
| 1 | Lymphocyte count mean or median | 0.927 | 0.828–1 | 15.3659 | 0.7010 |
| 2 | Neutrophil count mean or median | 0.903 | 0.784–1 | 17.4015 | 0.6450 |
| 3 | Age mean or median | 0.878 | 0.735–1 | 20.8519 | 0.5386 |
| 4 | White blood cell count mean or median | 0.868 | 0.715–1 | 20.6280 | 0.5460 |
| 5 | Age mean or median > 60.73 | 0.833 | 0.682–0.985 | 21.0499 | 0.5320 |
| Run 6: $m = 24$ | | | | | |
| 1 | Lymphocyte count mean or median | 0.927 | 0.828–1 | 15.3659 | 0.7010 |
| 2 | Neutrophil count mean or median | 0.903 | 0.784–1 | 17.4015 | 0.6450 |
| 3 | Age mean or median | 0.878 | 0.735–1 | 20.8519 | 0.5386 |
| 4 | Age mean or median > 60.72 | 0.833 | 0.682–0.985 | 21.0499 | 0.5320 |
| **Run 7: $m = 30$** |  |  |  |  |  |
| 1 | Lymphocyte count mean or median | 0.933 | 0.850–1 | 18.2773 | 0.7203 |
| 2 | Age mean or median | 0.829 | 0.668–0.990 | 31.0932 | 0.3936 |
| 3 | Age mean or median > 59.82 | 0.767 | 0.610–0.923 | 32.5430 | 0.3471 |

<sup>1</sup> The simple logistic regression models were listed in the order of the area under the curve (AUC) of Receiver Operating Characteristic (ROC).

**Table 2‒B:**
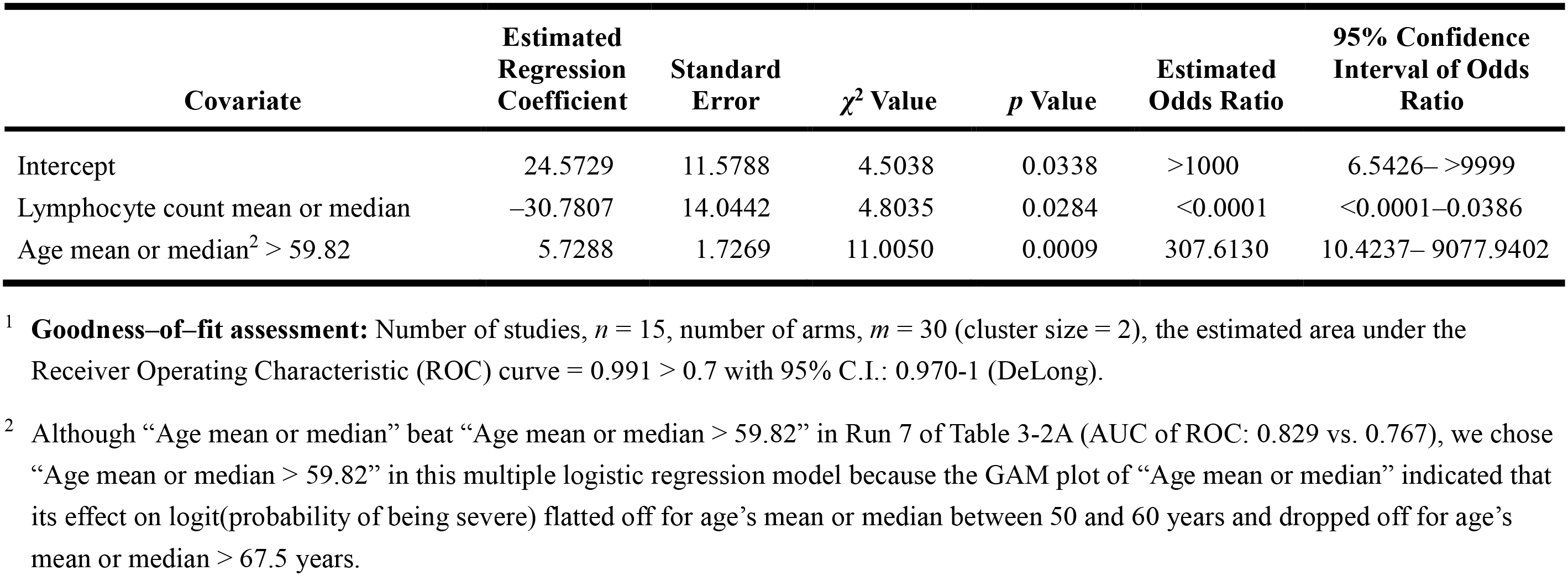
Multivariate analysis of the predictors for critical severity (critically severe vs. non-critically severe) by fitting a final multiple logistic regression model with the generalized estimating equations (GEE) method (assuming an ‘exchangeable’ working correlation structure).^1^

### Predictors for Mortality (Dead vs. Alive)

Results of univariate analyses of the predictors for mortality (dead vs. alive) were shown in Table 3-A. At the last run (seventh, *m* = 38) only lymphocyte count (AUC = 0.935) or lymphocyte count ≤ 0.87 (AUC = 0.895) and age (AUC = 0.913) or age > 67.28 (AUC = 0.895) existed in the final univariate analyses. Table 3-B listed the result of multivariate analysis for the predictors of mortality. Older age mean or median > 67.28 have a higher risk of mortality (HR = 17.3756, *p* = 0.0079) while lower lymphocyte count mean or median ≤ 0.87 had a higher risk of mortality (HR = 17.3756, *p* = 0.0079).

**Table 3-A:** Univariate analyses of the predictors for mortality (dead vs. alive) by fitting a series of simple logistic regression models with the generalized estimating equations (GEE) method (assuming an ‘exchangeable’ working correlation structure).

| Order<sup>1</sup> | Covariate | AUC of ROC | 95% C.I. of<br>AUC of ROC | Residual<br>Deviance | R<sup>2</sup> |
| --- | --- | --- | --- | --- | --- |
| **Run 1: $m = 12$** |  |  |  |  |  |
| 1 | Age mean or median | 1.000 | 1–1 | 0.0000 | 1.0000 |
| 2 | Lymphocyte count mean or median | 0.986 | 0.948–1 | 2.7726 | 0.9134 |
| 3 | CRP mean or median | 0.972 | 0.895–1 | 3.6475 | 0.8816 |
| 4 | Lactate dehydrogenase mean or median | 0.944 | 0.816–1 | 7.1707 | 0.7274 |
| 5 | Neutrophil count mean or median | 0.944 | 0.816–1 | 5.0828 | 0.8242 |
| 6 | White blood cell count mean or median | 0.944 | 0.816–1 | 6.2940 | 0.7701 |
| 7 | D-dimer mean or median | 0.917 | 0.738–1 | 8.4516 | 0.6592 |
| 8 | D-dimer mean or median > 1.94 | 0.917 | 0.753–1 | 5.7416 | 0.7955 |
| 9 | Lactate dehydrogenase mean or median > 443.26 | 0.917 | 0.753–1 | 5.7416 | 0.7955 |
| 10 | Platelet count mean or median | 0.861 | 0.625–1 | 9.9462 | 0.5698 |
| 11 | Platelet count mean or median $\leq 183.69$ | 0.833 | 0.602–1 | 10.8135 | 0.5125 |
| **Run 2: $m = 14$** |  |  |  |  |  |
| 1 | Lymphocyte count mean or median | 0.990 | 0.962–1 | 2.7726 | 0.9270 |
| 2 | Age mean or median | 0.959 | 0.869–1 | 6.3268 | 0.8096 |
| 3 | Neutrophil count mean or median | 0.959 | 0.864–1 | 5.1213 | 0.8528 |
| 4 | White blood cell count mean or median | 0.959 | 0.864–1 | 6.3653 | 0.8081 |
| 5 | Lactate dehydrogenase mean or median | 0.939 | 0.806–1 | 10.8264 | 0.6110 |
| 6 | CRP mean or median | 0.878 | 0.667–1 | 13.2424 | 0.4750 |
| 7 | CRP mean or median > 85.76 | 0.857 | 0.659–1 | 11.4833 | 0.5763 |
| 8 | Lactate dehydrogenase mean or median > 503.93 | 0.857 | 0.659–1 | 11.4833 | 0.5763 |
| **Run 3: $m = 16$** |  |  |  |  |  |
| 1 | Lymphocyte count mean or median | 0.992 | 0.971–1 | 2.7726 | 0.9369 |
| 2 | Lactate dehydrogenase mean or median | 0.938 | 0.807–1 | 12.4176 | 0.6090 |
| 3 | Age mean or median | 0.922 | 0.794–1 | 10.8942 | 0.6748 |
| 4 | Neutrophil count mean or median | 0.906 | 0.756–1 | 10.2175 | 0.7021 |
| 5 | White blood cell count mean or median | 0.906 | 0.756–1 | 11.8361 | 0.6349 |
| 6 | Lactate dehydrogenase mean or median > 501.70 | 0.875 | 0.702–1 | 12.0566 | 0.6252 |
| 7 | White blood cell count mean or median > 6.74 | 0.812 | 0.611–1 | 15.2763 | 0.4673 |
| **Run 4: $m = 20$** |  |  |  |  |  |
| 1 | Lymphocyte count mean or median | 0.990 | 0.966–1 | 3.8191 | 0.9299 |
| 2 | Age mean or median | 0.930 | 0.821–1 | 12.1678 | 0.7208 |
| 3 | Neutrophil count mean or median | 0.920 | 0.797–1 | 12.6233 | 0.7067 |
| 4 | Lactate dehydrogenase mean or median | 0.910 | 0.783–1 | 16.9125 | 0.5569 |
| 5 | Lactate dehydrogenase mean or median > 479.84 | 0.850 | 0.687–1 | 16.7100 | 0.5647 |
| **Run 5: $m = 28$** |  |  |  |  |  |
| 1 | Lymphocyte count mean or median | 0.995 | 0.983–1 | 3.8191 | 0.9513 |
| 2 | Age mean or median | 0.954 | 0.882–1 | 13.6223 | 0.7911 |
| 3 | Neutrophil count mean or median | 0.913 | 0.811–1 | 19.5828 | 0.6625 |
| **Run 6: $m = 38$** |  |  |  |  |  |
| 1 | Lymphocyte count mean or median | 0.935 | 0.836–1 | 31.0924 | 0.5778 |
| 2 | Age mean or median | 0.913 | 0.807–1 | 30.6503 | 0.5866 |
| 3 | Age mean or median > 67.28 | 0.895 | 0.796–0.994 | 24.8313 | 0.6926 |
| 4 | Lymphocyte count mean or median ≤ 0.87 | 0.895 | 0.796–0.994 | 24.8313 | 0.6926 |

<sup>1</sup> The simple logistic regression models were listed in the order of the area under the curve (AUC) of Receiver Operating Characteristic (ROC).

**Table 3-B:** Multivariate analysis of the predictors for mortality (dead vs. alive) by fitting a final multiple logistic regression model with the generalized estimating equations (GEE) method (assuming an ‘independence’ working correlation structure).^1^

| Covariate | Estimated Regression Coefficient | Standard Error | $\chi^2$ Value | *p* Value | Estimated Odds Ratio | 95% Confidence Interval of Odds Ratio |
| --- | --- | --- | --- | --- | --- | --- |
| Intercept | -2.8551 | 1.0052 | 17.5966 | <0.0001 | 0.0576 | 0.0045–0.2744 |
| Lymphocyte count mean or median $\leq 0.87$ | 2.8551 | 1.2055 | 7.0560 | 0.0079 | 17.3756 | 2.1157–232.7834 |
| Age mean or median<sup>2</sup> $> 67.28$ | 2.8551 | 1.2055 | 7.0560 | 0.0079 | 17.3756 | 2.1157–232.7834 |

<sup>1</sup> **Goodness-of-fit assessment:** Number of studies, $n = 19$ , number of arms, $m = 38$ (cluster size = 2), the estimated area under the Receiver Operating Characteristic (ROC) curve = $0.954 > 0.7$ with 95% C.I.: 0.888-1 (DeLong). Yet, the regression coefficients and the standard errors were estimated by fitting the corresponding Firth’s bias-reduced logistic regression model with the `logistf` function of the `logistf` package in R owing to the separation (or high discrimination) problem.
<sup>2</sup> Although “Age mean or median” beat “Age mean or median $> 67.28$ ” in Run 6 of Table 3-3A (AUC of ROC: 0.913 vs. 0.895), we chose “Age mean or median $> 67.28$ ” in this multiple logistic regression model because the GAM plot of “Age mean or median” indicated that its effect on $\text{logit}(\text{probability of being dead})$ dropped off for age’s mean or median $> 77.5$ years.

## Discussion

The results of this study provide numerous imperative insights. After comparisons lymphocyte count was the most powerful predictor among the fourteen explored lab items. Single lab data, lymphocyte count at initial hospital presentation together with age can be remarkable indicators to discriminate the health status (severe vs. non-severe, critically severe vs. non-critically severe) or the final consequence (dead vs. alive) for COVID-19 patients. Compared with vital signs, symptoms, comorbidities, several lab data (CRP, D-dimer, lymphocyte, neutrophil, platelet, LDH) holds the value to differentiate disease severity and to predict the mortality of COVID-19 patients. To the best of our knowledge, this was the first meta-analysis study that potential bias of subject duplication of COVID-19 patients in studies has been eliminated before analyses.

After SARS-CoV2 infection, multiple mechanisms of the human body triggered, e.g., immune (ex. WBC, lymphocyte, neutrophil) responses, inflammatory cataracts (ex. CRP, PCT), and the activation of coagulation cascades (ex. platelet count, D-dimer).^108–111^ After virus invasion to the tissues which starts early, the inflammation situation intensifies^110, 112^, the inflammatory indicators will increase dramatically.^15, 28, 29, 113, 114^ The wide distribution of the COVID-19 receptors, e.g., angiotensin-converting enzyme-2 (ACE2) receptors, abundantly expressed in a variety of cells residing in many human organs, could exaggerate systemic failure due to direct organ injury.^115, 116^ Organ (ex. lung, liver, kidney, heart, brain, etc.) damage indicators such as ALT, AST, total bilirubin, LDH, hypersensitive troponin I, etc. will be augmented to reflecting the impairment situation.^30, 117, 118^ Our study confirmed again that levels of several laboratory data although not all are profound predictors to disease severity or mortality for COVID-19 patients as compared with previous studies.^15, 28, 29, 30, 114^

It is no surprise that lymphocyte count played such an important role to COVID-19 patients in defending SARS-CoV-2.^28, 109, 112^ Adaptive immune cells such as lymphocytes are essential for virus clearance as well as for recovery from the disease.^109, 112, 119^ Interaction between SARS-CoV-2 and the immune system of an individual results in a diverse clinical manifestation.^16, 75, 88, 94, 96, 112^ Our study reveals that lymphocyte count offerings a defensive feature to COVID-19 patients within a certain range (Table 1-B, Table 3-B). Lower lever (e.g., lymphocyte count ≤ 1.03) or extreme lower (e.g., lymphocyte count ≤ 0.87) implies immune weakness and worsens outcomes to disease severity or mortality. However, a too high level of immune response becomes another issue, which may induce unintended results such as cytokine storm.^75, 88^ In our study, a higher level of lymphocyte count (e.g., lymphocyte count > 2.06) revealed a more severe status to severity (Table 1-B). A current hypothesis is that a cytokine storm can induce or further aggravate SARS-CoV-2 infection.^120, 121^ The degree to which SARS-CoV-2 targets immune cells remains poorly defined. It is crucial to understand more about the interaction of SARS-CoV-2 with the host immune system and the subsequent contribution to the organ functions and disease progression.

Plentiful tools, e.g., demographics, symptoms, vital signs, comorbidities, imaging, etc., have been explored to their prediction capability for COVID-19 patients.^28, 78, 114^ However, such data has its limitations. Routine lab data retains several advantages, which can indicate the whole body situation of a COVID-19 patient whose functions can be changed dramatically in few days.^122^ Additionally lab testing is easy to access, repeatable, self-explain, relatively cheap, and therefore can be a cost-effective tool under pandemic circumstances.

Current criteria to judge the severity, to triage or referral COVID-19 patients, are based on imaging, demographics, comorbidities, vital signs, or symptoms.^6, 123, 124^ Based on our study results single lab data, lymphocyte count at administration plus age can be useful for the purposes. Early and continue monitoring lab data for a COVID-19 patient can help to understand the health state, triage the patient, predict the severity of disease, predict the health consequences, and workout treatment judgment appropriately.

Our study has several limitations. Due to the lack of non-English articles, pediatric study and specific disease groups, interpretation of the results must be cautious. Ideally, all desired lab data should be collected and analyzed in all studies. However, it is not realistic in the real world because of wide-ranging medical resource deficiency that existed across countries. We suggest collecting essential data through a standardized list while clinical presentation, medical history, imaging information, comprehensive lab data, and other valuable factors, can be assembled and analyzed which will accelerate knowledge accumulation in particular under global pandemics. Retrospective observational study conducted at the level of hospital or community, characteristics of individual patients could not be retrieved. In addition, the dynamic relationship among various lab data, the status of disease progression, functions, and feelings of the patient, have not been explored due to inadequate data. More extensive and large-scale studies are required to double confirm the findings of this study.

## Conclusion

Our study involved 26,627 confirmed COVID-19 patients across sixteen countries provides evidence for defending disease under pandemics. Results prove that lymphocyte count is a universal biomarker to disease severity and mortality across regions. Several routine lab data at the initial hospital presented good prediction capability. Routine lab testing could be a useful tool in particular under a pandemic condition whereas medical resource is constrained. How to maintain or improve good immunity levels for the general population in daily life can be a crucial strategy to stakeholders in facing life-threatening infectious diseases such as COVID-19.

## Data Availability

The data of this study were available from the corresponding author on request.

## Contributors

KLL and FCH designed the study and take responsibility for the integrity of the data and the accuracy of the data analysis. JJC and KLL were in charge of the systematic review and data collection. FCH and FYW conducted the statistical analysis. KLL and FGH contributed to the writing of the manuscript. All authors contributed to data interpretation, reviewed, and approved the final version.

## Data sharing

The data of this study were available from the corresponding author on request.

## Acknowledgments

None.

## Ethical statement

The study was a systematic review and meta-regression analysis so that ethical approval was not needed.

## Supplementary material

Supplementary data to this article can be found online at https://.

---

**Appendix I:** The profile of the collected 76 studies with a total of 26,627 laboratory-confirmed COVID-19 patients.^1^

| Study | Country | Recruitment Period<sup>2</sup> | Total | Non-Severe | Severe | Non-Critical Severe | Critical Severe | Alive | Dead |
| --- | --- | --- | --- | --- | --- | --- | --- | --- | --- |
| Almazeedi Sulaiman et al (41) | Kuwait | Feb 24-Apr 20 | 1096 | – | – | 1054 | 42 | 1077 | 19 |
| Bahl Amit et al (24) | USA | Mar 1-Mar 31 | 1461 | – | – | – | – | 1134 | 327 |
| Berenguer Juan et al (42) | Spain | Feb-Mar 17 | 4035 | – | – | – | – | 2904 | 1131 |
| Bonetti Graziella et al (25) | Italy | Mar 1-Mar 30 | 144 | – | – | – | – | 74 | 70 |
| Botero Diana Maria Ronderos et al (43) | USA | Mar 15-Apr 23 | 157 | – | – | – | – | 132 | 25 |
| Brill Simon E. et al (44) | UK | Mar 10-Apr 8 | 410 | – | – | – | – | 237 | 173 |
| Cai Qingxian et al (45) | China | Jan 11-Feb 6 | 298 | 240 | 58 | – | – | – | – |
| Chen Lei et al (18) | China | Jan 20-Apr 4 | 1859 | – | – | – | – | 1651 | 208 |
| Chen Tao et al (46) | China | Jan 13-Feb 12 | 274 | – | – | – | – | 161 | 113 |
| Ciceri Fabio et al (23) | Italy | Feb 25-Mar 24 | 410 | – | – | – | – | 315 | 95 |
| Cipriani Alberto et al (47) | Italy | Feb 26-Mar 31 | 109 | – | – | – | – | 89 | 20 |
| Fan Bingwen Eugene et al (48) | Singapore | Jan 23-Feb 4 | 67 | – | – | 58 | 9 | – | – |
| Ferguson Jessica et al (49) | USA | Mar 13-Apr 11 | 72 | – | – | 51 | 21 | – | – |
| Gao Yong et al (50) | China | Jan 23-Feb 2 | 43 | 28 | 15 | – | – | – | – |
| Gayam Vijay et al (51) | USA | Mar 1-Apr 9 | 408 | – | – | – | – | 276 | 132 |
| Ghweil Ali A et al (10) | Egypt | Mar-Apr | 66 | 36 | 30 | – | – | – | – |
| Giacomelli Andrea et al (52) | Italy | Feb 21-Mar 19 | 233 | – | – | – | – | 185 | 48 |
| He Li et al (53) | China | Feb 10-Apr 9 | 174 | 120 | 54 | – | – | – | – |
| Hong Kyung Soo et al (54) | South Korea | – | 98 | – | – | 85 | 13 | – | – |
| Huang Chaolin et al (55) | China | Dec 16 '19-Jan 2 | 41 | – | – | 28 | 13 | – | – |
| Huang Huihuang et al (17) | China | Jan 13-Mar 10 | 64 | 43 | 21 | – | – | – | – |
| Israelsen Simone Bastrup et al (56) | Denmark | Mar 10-Apr 23 | 175 | – | – | 148 | 27 | – | – |
| Javannian Mostafa et al (57) | Iran | Feb 25-Mar 12 | 100 | – | – | – | – | 81 | 19 |
| Jiang Hao et al (58) | China | Feb-Mar | 59 | – | – | 15 | 44 | – | – |
| Khalil K. et al (59) | UK | Mar 7-Apr 7 | 220 | – | – | – | – | 162 | 58 |
| Khamis Faryal et al (60) | Oman | Feb 24-Apr 24 | 63 | – | – | 39 | 24 | – | – |
| Lee Ji Yeon et al (61) | South Korea | Feb 18-Mar 4 | 98 | – | – | – | – | 78 | 20 |
| Lee Ji Yeon et al (62) | South Korea | Feb 21-Apr 2 | 694 | 557 | 137 | – | – | – | – |
| Lendorf Maria Elisabeth et al (63) | Denmark | Mar 1-May 4 | 111 | – | – | 91 | 20 | – | – |
| Li Huan et al (64) | China | Jan 18-Feb 26 | 132 | 60 | 72 | 116 | 16 | – | – |
| Li Kunhua et al (65) | China | Jan -Feb | 83 | 58 | 25 | – | – | – | – |
| Li Tao et al (66) | China | Feb 1-Mar 31 | 312 | 207 | 105 | – | – | – | – |
| Li Xiaolei et al (67) | China | Jan 24-Mar 12 | 215 | 159 | 56 | – | – | – | – |
| Liu Jiao et al (68) | China | Dec 29 '19-Feb 28 | 1190 | – | – | – | – | 1033 | 157 |
| Liu Songqiao et al (69) | China | Jan 10-Mar 15 | 625 | 561 | 64 | – | – | – | – |
| Lv Zhihua et al (70) | China | Feb 4-Feb 28 | 354 | 115 | 239 | 270 | 84 | – | – |
| Mao Ling et al (71) | China | Jan 16-Feb 19 | 214 | 126 | 88 | – | – | – | – |
| Ortiz-Brizuela Edgar et al (72) | Mexico | Feb 26-Apr 11 | 140 | – | – | 111 | 29 | – | – |
| Qian Guo-Qing et al (73) | China | Jan 20-Feb 11 | 91 | 82 | 9 | – | – | – | – |
| Qun Sen et al (16) | China | Feb 2-Mar 17 | 230 | – | – | 225 | 5 | – | – |
| Romero-Sanchez Carlos M. et al (74) | Spain | Mar 1-Apr 1 | 841 | 512 | 329 | – | – | – | – |
| Satış Hasan et al (75) | Turkey | – | 38 | 27 | 11 | – | – | – | – |
| Shahriarirad Reza et al (76) | Iran | Feb 20-Mar 20 | 113 | – | – | 102 | 11 | 104 | 9 |
| Shang Weifeng et al (77) | China | Jan 16-Feb 28 | 443 | 304 | 139 | – | – | – | – |
| Shu Zixin et al (21) | China | Jan 15-Mar 2 | 293 | 207 | 86 | – | – | – | – |
| Sun Dong et al (78) | China | Jan 23-Feb 19 | 84 | 61 | 23 | – | – | – | – |
| Sun Haiying et al (19) | China | Jan 29-Mar 5 | 244 | – | – | – | – | 123 | 121 |
| Sun Suyu et al (79) | China | Jan 19-Feb 20 | 116 | 89 | 27 | – | – | – | – |
| Sun Ying et al (80) | China | Jan-Apr | 63 | – | – | 54 | 9 | – | – |
| Taghiloo Saeid et al (81) | Iran | Mar-Apr | 61 | 39 | 22 | – | – | – | – |
| Tao Zheyang et al (82) | China | Dec 1 ‘19-Mar | 222 | 202 | 20 | – | – | – | – |
| Uchida Yoshihito et al (83) | Japan | Mar-Jun | 35 | 27 | 8 | – | – | – | – |
| Vena Antonio et al (84) | Italy | Feb 25-Mar 25 | 275 | – | – | – | – | 155 | 120 |
| Wang Changzheng et al (85) | China | Jan 23-Feb 13 | 161 | 131 | 30 | – | – | – | – |
| Wang Dan et al (86) | China | Jan 15-Feb 28 | 143 | 72 | 71 | – | – | – | – |
| Wang Dawei et al (87) | China | Jan 1-Jan 28 | 138 | – | – | 102 | 36 | – | – |
| Wang Feng et al (88) | China | – | 65 | – | – | 50 | 15 | – | – |
| Wang Ruirui et al (89) | China | Jan 22-Feb 18 | 125 | 100 | 25 | – | – | – | – |
| Wang Ruoran et al (90) | China | Jan 30-Feb 24 | 450 | – | – | – | – | 372 | 78 |
| Wu J. et al (91) | China | Jan 20-Feb 19 | 280 | 197 | 83 | – | – | – | – |
| Yan Xiquan et al (92) | China | Jan 21-Jun 27 | 194 | 156 | 38 | 180 | 14 | – | – |
| Yang Luhuan et al (93) | China | Jan 30-Feb 8 | 200 | – | – | 171 | 29 | – | – |
| Yang Miao et al (94) | China | Jan 17-Mar 12 | 108 | 84 | 24 | – | – | – | – |
| Yao Qingchun et al (95) | China | Jan 30-Feb 11 | 96 | 83 | 13 | – | – | – | – |
| Ye Wenjing et al (96) | China | Jan 1-Mar 16 | 349 | – | – | – | – | 297 | 52 |
| Yu Caizheng et al (97) | China | Jan 14-Feb 28 | 1663 | 799 | 864 | – | – | – | – |
| Zerah Lorene et al (98) | France | Mar 13-Apr 15 | 821 | – | – | – | – | 571 | 250 |
| Zhang Chi et al (99) | China | Jan-Feb | 80 | 56 | 21 | – | – | 77 | 3 |
| Zhang Jin-jin et al (100) | China | Jan 16-Feb 3 | 140 | 82 | 58 | – | – | – | – |
| Zhang Jin-jin et al (101) | China | Dec 29 ‘19-Feb 16 | 289 | – | – | – | – | 240 | 49 |
| Zhang Shan-Yan et al (22) | China | Jan 17-Feb 12 | 788 | 710 | 78 | 771 | 17 | – | – |
| Zhang Wenjing et al (14) | China | Jan-Mar | 90 | 70 | 20 | – | – | – | – |
| Zhao Zirun et al (102) | USA | Mar 9-Apr 20 | 593 | – | – | 398 | 195 | – | – |
| Zheng Fang et al (103) | China | Jan 17-Feb 7 | 161 | 131 | 30 | – | – | – | – |
| Zheng Yongli et al (104) | China | Jan 16-Feb 20 | 99 | 67 | 32 | – | – | – | – |
| Zheng Yufen et al (20) | China | Jan 17-Feb 26 | 141 | 112 | 29 | – | – | – | – |
| **Sub-total** |  |  | – | **9,764** |  | **4,792** |  | **14,825** |  |
| **Total** |  |  | **26,627** | **6,710** | **3,054** | **4,119** | **673** | **11,528** | **3,297** |

<sup>1</sup> The detailed data can be found in the Excel file in the online supplementary materials.
<sup>2</sup> The subjects were recruited during the year 2020 if not specified.

**Appendix II-1:** The summary statistics of the demographics, clinical characteristics, comorbidities, and laboratory data of the COVID-19 patients on initial hospital presentations from the collected 38 studies for the assessment of severity.

| Variable | Number of<br>Studies<sup>1</sup><br>(*n*) | Non-Severe |  |  | Severe |  |  | *p* Value<sup>2</sup> |
| --- | --- | --- | --- | --- | --- | --- | --- | --- |
|  |  | Median | Minimum | Maximum | Median | Minimum | Maximum |  |
| Demographics |  |  |  |  |  |  |  |  |
| Age (years) | 38 | 46.50 | 37.55 | 67.10 | 62.55 | 39.47 | 75.00 | <0.0001 |
| Male (%) | 35 | 50.40 | 27.80 | 77.80 | 60.00 | 38.10 | 75.00 | 0.0018 |
| Current smoker (%) | 7 | 6.58 | 0 | 36.10 | 8.98 | 0 | 30.00 | 0.5649 |
| Vital signs |  |  |  |  |  |  |  |  |
| Heart rate (beats/min) | 4 | 86.42 | 82.00 | 89.00 | 89.99 | 86.00 | 92.00 | 0.1465 |
| Respiratory rate (breaths/min) | 3 | 20.00 | 18.87 | 20.00 | 20.98 | 20.00 | 21.00 | 0.1642 |
| Systolic pressure (mmHg) | 2 | 108.15 | 93.30 | 123.00 | 111.95 | 91.00 | 132.90 | 1 |
| Clinical features |  |  |  |  |  |  |  |  |
| Fever (%) | 26 | 73.36 | 16.80 | 97.20 | 89.75 | 38.30 | 100 | 0.0030 |
| Cough (%) | 26 | 58.97 | 31.50 | 81.50 | 69.60 | 15.00 | 100 | 0.0570 |
| Fatigue (%) | 17 | 25.71 | 4.17 | 76.10 | 40.00 | 5.17 | 87.50 | 0.3394 |
| Myalgia (%) | 15 | 14.21 | 2.00 | 43.50 | 23.53 | 0 | 51.81 | 0.2290 |
| Dyspnea (%) | 11 | 11.90 | 1.65 | 44.40 | 30.00 | 2.15 | 80.00 | 0.0417 |
| Sputum production (%) | 14 | 23.70 | 6.90 | 88.10 | 37.55 | 9.00 | 96.20 | 0.1636 |
| Headache (%) | 17 | 10.30 | 1.20 | 25.60 | 13.30 | 0 | 38.55 | 0.4484 |
| Sore throat (%) | 12 | 14.11 | 0.83 | 26.10 | 10.84 | 0 | 46.70 | 0.7949 |
| Diarrhea (%) | 19 | 8.21 | 0.51 | 28.20 | 7.62 | 0 | 50.00 | 0.9070 |
| Shortness of breath (%) | 5 | 22.86 | 6.90 | 45.80 | 25.00 | 8.00 | 100 | 0.5476 |
| Chest congestion / Chest tightness (%) | 5 | 29.00 | 4.50 | 40.30 | 45.30 | 0 | 50.00 | 0.4206 |
| Chest pain (%) | 6 | 2.10 | 1.02 | 15.80 | 13.42 | 2.15 | 20.00 | 0.0931 |
| **Comorbidities** |  |  |  |  |  |  |  |  |
| Any (%) | 14 | 32.10 | 6.90 | 70.50 | 58.27 | 31.38 | 90.50 | 0.0033 |
| Hypertension (%) | 27 | 14.80 | 2.90 | 48.00 | 37.90 | 4.30 | 82.90 | <0.0001 |
| Diabetes (%) | 28 | 8.30 | 0 | 34.80 | 16.30 | 6.70 | 46.70 | <0.0001 |
| Cardiovascular disease (%) | 26 | 4.68 | 0 | 22.70 | 12.75 | 0 | 47.37 | 0.0014 |
| Chronic obstructive pulmonary disease (%) | 21 | 1.74 | 0 | 7.73 | 6.70 | 0 | 24.00 | 0.0003 |
| Malignancy (%) | 16 | 1.63 | 0 | 7.40 | 4.46 | 0 | 10.30 | 0.0328 |
| Cerebrovascular disease (%) | 11 | 2.80 | 0.94 | 7.50 | 7.90 | 0 | 25.91 | 0.0086 |
| Chronic liver disease (%) | 15 | 3.10 | 0 | 8.33 | 4.50 | 0 | 20.80 | 0.2444 |
| Chronic renal disease (%) | 16 | 0.84 | 0 | 6.70 | 4.61 | 1.70 | 13.00 | 0.0006 |
| **Blood routine** |  |  |  |  |  |  |  |  |
| White blood cell count ($\times 10^9/L$) | 36 | 5.00 | 3.60 | 8.59 | 5.75 | 3.40 | 12.10 | 0.0047 |
| Neutrophil count ($\times 10^9/L$) | 28 | 3.22 | 2.40 | 6.69 | 4.43 | 1.65 | 10.51 | 0.0011 |
| Lymphocyte count ($\times 10^9/L$) | 36 | 1.30 | 0.80 | 1.90 | 0.81 | 0.48 | 2.19 | <0.0001 |
| Neutrophil/Lymphocyte ratio (NLR) | 4 | 2.49 | 2.38 | 7.93 | 8.15 | 4.75 | 29.90 | 0.1143 |
| Platelet count ($\times 10^9/L$) | 26 | 206.00 | 156.00 | 240.46 | 176.00 | 86.00 | 265.00 | 0.0077 |
| **Blood biochemistry** |  |  |  |  |  |  |  |  |
| Alanine aminotransferase (ALT) (U/L) | 25 | 23.00 | 18.00 | 40.53 | 28.54 | 19.90 | 54.00 | 0.0016 |
| Aspartate aminotransferase (AST) (U/L) | 23 | 25.00 | 16.00 | 42.60 | 34.00 | 26.00 | 61.00 | 0.0003 |
| Total bilirubin ($\mu\text{mol/L}$) | 15 | 9.80 | 6.60 | 13.70 | 11.30 | 6.70 | 21.20 | 0.0128 |
| Lactate dehydrogenase (LDH) (U/L) | 17 | 209.20 | 150.40 | 489.00 | 287.00 | 220.10 | 695.19 | 0.0001 |
| **Coagulation function** |  |  |  |  |  |  |  |  |
| D-dimer (mg/L or $\mu\text{g/mL}$) | 18 | 0.40 | 0.20 | 1.90 | 0.93 | 0.30 | 7.93 | 0.0011 |

| **Inflammatory markers** |  |  |  |  |  |  |  |  |
| --- | --- | --- | --- | --- | --- | --- | --- | --- |
| C-reactive protein (CRP) (mg/L) | 29 | 9.70 | 1.70 | 106.66 | 53.80 | 19.50 | 291.00 | <0.0001 |
| Procalcitonin (PCT) (×100 ng/mL) | 22 | 4.65 | 0.13 | 162.00 | 1.00 | 0 | 331.00 | 0.0280 |
| **Myocardial injury marker** |  |  |  |  |  |  |  |  |
| Hypersensitive troponin I (hs-cTnI) (×100 ng/mL) | 3 | 0.75 | 0.40 | 1.00 | 1.53 | 0.70 | 5.00 | 0.4000 |

<sup>1</sup> It was the number of studies without missing values among the collected 38 studies.
<sup>2</sup> The listed *p* values were calculated using the Wilcoxon rank-sum test based on the available number of studies.

**Appendix II-2:** The summary statistics of the demographics, clinical characteristics, and biomarkers of the COVID-19 patients on initial hospital presentations from the collected 21 studies for the assessment of critical severity.

| Variable | Number of<br>Studies<sup>1</sup><br>(*n*) | Non-Critically Severe |  |  | Critically Severe |  |  | *p* Value<sup>2</sup> |
| --- | --- | --- | --- | --- | --- | --- | --- | --- |
|  |  | Median | Minimum | Maximum | Median | Minimum | Maximum |  |
| Demographics |  |  |  |  |  |  |  |  |
| Age (years) | 19 | 52.00 | 37.10 | 73.00 | 63.20 | 49.00 | 71.00 | 0.0030 |
| Male (%) | 18 | 53.78 | 37.60 | 82.00 | 63.80 | 46.20 | 88.00 | 0.0095 |
| Current smoker (%) | 5 | 10.96 | 6.71 | 21.90 | 11.80 | 0 | 26.70 | 1 |
| Vital signs |  |  |  |  |  |  |  |  |
| Heart rate (beats/min) | 5 | 87.00 | 84.90 | 98.00 | 89.00 | 83.50 | 100 | 0.6742 |
| Respiratory rate (breaths/min) | 6 | 20.45 | 20.00 | 22.00 | 22.50 | 20.00 | 30.00 | 0.1832 |
| Systolic pressure (mmHg) | 6 | 125.15 | 90.00 | 130.00 | 126.05 | 91.00 | 145.00 | 0.6304 |
| Clinical features |  |  |  |  |  |  |  |  |
| Fever (%) | 15 | 73.93 | 51.00 | 98.00 | 77.80 | 45.00 | 100 | 0.6630 |
| Cough (%) | 15 | 64.03 | 27.60 | 90.10 | 66.70 | 45.00 | 100 | 1 |
| Fatigue (%) | 11 | 26.00 | 3.30 | 65.70 | 35.30 | 7.10 | 90.90 | 0.4779 |
| Myalgia (%) | 14 | 24.95 | 6.70 | 75.50 | 25.25 | 9.00 | 79.30 | 0.9100 |
| Dyspnea (%) | 8 | 35.00 | 4.10 | 61.50 | 73.40 | 20.00 | 92.00 | 0.0086 |
| Sputum production (%) | 10 | 25.25 | 1.70 | 48.74 | 20.20 | 9.70 | 86.00 | 0.9698 |
| Headache (%) | 13 | 14.60 | 5.90 | 80.18 | 9.00 | 0 | 62.07 | 0.3048 |
| Sore throat (%) | 10 | 14.42 | 9.50 | 32.40 | 11.75 | 0 | 33.30 | 0.3642 |
| Diarrhea (%) | 12 | 9.01 | 1.40 | 33.30 | 10.15 | 0 | 37.90 | 0.6441 |
| Shortness of breath (%) | 7 | 21.00 | 1.70 | 74.00 | 66.67 | 28.60 | 95.00 | 0.0530 |
| Chest congestion / Chest tightness (%) | 1 | 15.80 | 15.80 | 15.80 | 15.40 | 15.40 | 15.40 | 1 |
| Chest pain (%) | 3 | 33.30 | 11.80 | 36.30 | 54.50 | 9.50 | 55.20 | 0.7000 |
| **Comorbidities** |  |  |  |  |  |  |  |  |
| Any (%) | 8 | 31.12 | 26.22 | 67.00 | 58.75 | 33.33 | 100 | 0.0104 |
| Hypertension (%) | 14 | 25.50 | 14.00 | 42.70 | 40.60 | 15.00 | 88.20 | 0.0094 |
| Diabetes (%) | 15 | 12.70 | 5.90 | 26.10 | 23.50 | 8.00 | 46.00 | 0.0128 |
| Cardiovascular disease (%) | 16 | 11.30 | 1.32 | 51.40 | 17.00 | 0 | 60.00 | 0.2351 |
| Chronic obstructive pulmonary disease (%) | 14 | 2.68 | 0 | 9.00 | 5.75 | 0 | 36.00 | 0.1901 |
| Malignancy (%) | 12 | 1.90 | 0.74 | 8.00 | 1.20 | 0 | 11.10 | 0.4322 |
| Cerebrovascular disease (%) | 5 | 1.34 | 0.57 | 12.00 | 10.00 | 0 | 21.00 | 0.3457 |
| Chronic liver disease (%) | 10 | 2.15 | 0 | 6.74 | 1.20 | 0 | 11.80 | 0.6456 |
| Chronic renal disease (%) | 10 | 2.55 | 0.60 | 7.70 | 7.00 | 0 | 15.00 | 0.0638 |
| **Blood routine** |  |  |  |  |  |  |  |  |
| White blood cell count ($\times 10^9/L$) | 20 | 5.74 | 4.30 | 6.77 | 6.82 | 5.10 | 11.30 | 0.0001 |
| Neutrophil count ($\times 10^9/L$) | 13 | 3.21 | 1.70 | 4.64 | 5.54 | 3.50 | 10.60 | 0.0009 |
| Lymphocyte count ($\times 10^9/L$) | 17 | 1.14 | 0.86 | 1.60 | 0.80 | 0.40 | 1.37 | 0.0001 |
| Neutrophil/Lymphocyte ratio (NLR) | 1 | 2.50 | 2.50 | 2.50 | 6.32 | 6.32 | 6.32 | 1 |
| Platelet count ($\times 10^9/L$) | 15 | 193.00 | 149.00 | 245.00 | 196.00 | 142.00 | 253.00 | 0.9504 |
| **Blood biochemistry** |  |  |  |  |  |  |  |  |
| Alanine aminotransferase (ALT) (U/L) | 15 | 30.10 | 23.00 | 73.02 | 37.80 | 24.76 | 89.10 | 0.0084 |
| Aspartate aminotransferase (AST) (U/L) | 11 | 31.00 | 25.04 | 47.00 | 49.00 | 26.50 | 100.30 | 0.0104 |
| Total bilirubin ($\mu\text{mol/L}$) | 11 | 9.55 | 0.69 | 13.68 | 13.00 | 0.64 | 19.50 | 0.0612 |
| Lactate dehydrogenase (LDH) (U/L) | 15 | 281.00 | 210.41 | 686.23 | 443.50 | 320.50 | 1684.00 | 0.0009 |
| **Coagulation function** |  |  |  |  |  |  |  |  |
| D-dimer (mg/L or $\mu\text{g/mL}$) | 13 | 0.50 | <0.01 | 4.10 | 1.76 | 0.36 | 11.97 | 0.0333 |

| **Inflammatory markers** |  |  |  |  |  |  |  |  |
| --- | --- | --- | --- | --- | --- | --- | --- | --- |
| C-reactive protein (CRP) (mg/L) | 16 | 39.32 | 8.00 | 68.88 | 103.22 | 9.90 | 177.00 | 0.0005 |
| Procalcitonin (PCT) ($\times 100$ ng/mL) | 9 | 13.00 | 5.00 | 41.00 | 44.00 | 10.00 | 270.00 | 0.0331 |
| **Myocardial injury marker** |  |  |  |  |  |  |  |  |
| Hypersensitive troponin I (hs-cTnI) ($\times 100$ ng/mL) | 4 | 0.46 | 0.35 | 1.00 | 1.03 | 0.33 | 1.10 | 0.3836 |

<sup>1</sup> It was the number of studies without missing values among the collected 21 studies.
<sup>2</sup> The listed *p* values were calculated using the Wilcoxon rank-sum test based on the available number of studies.

**Appendix II-3:**
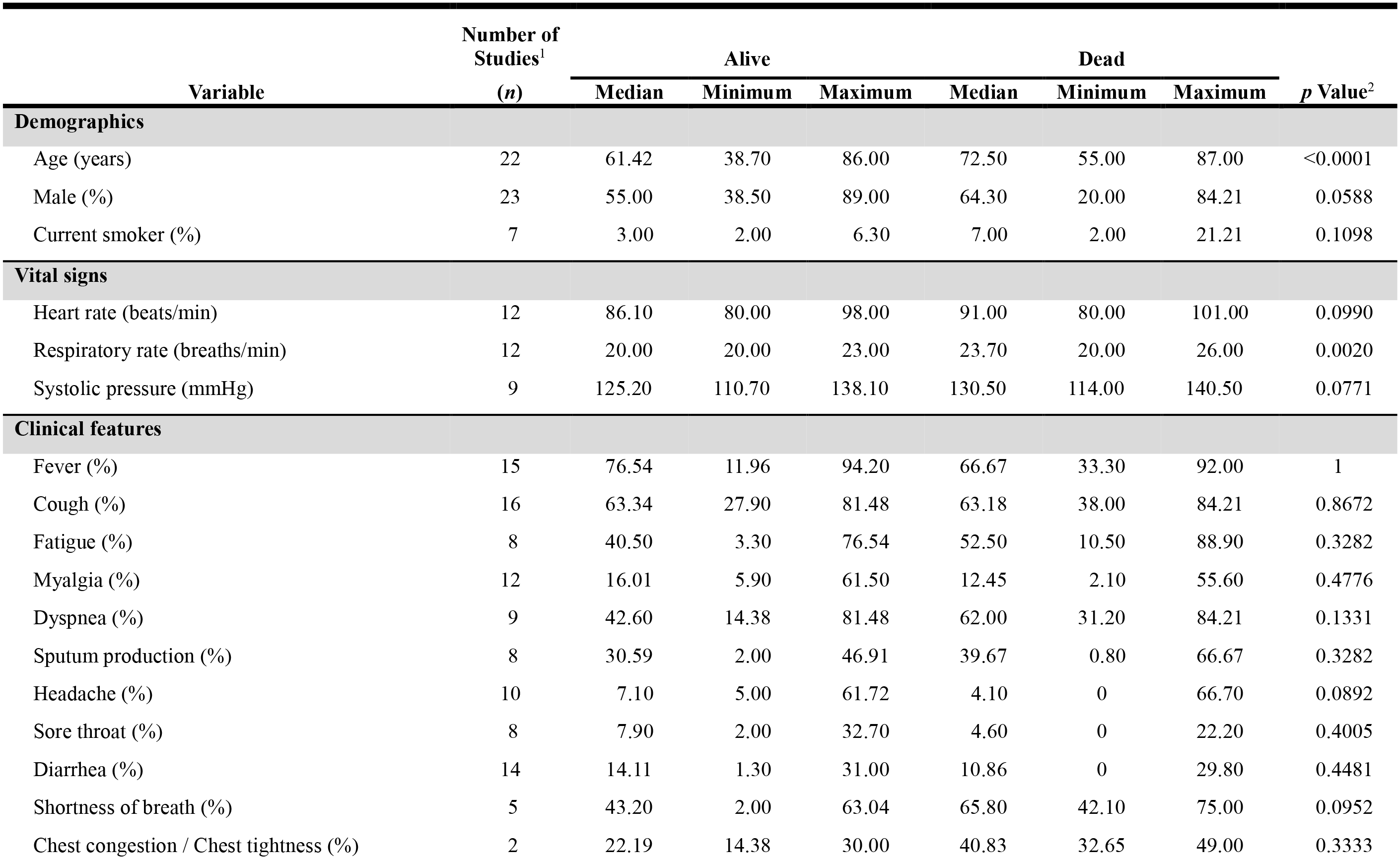
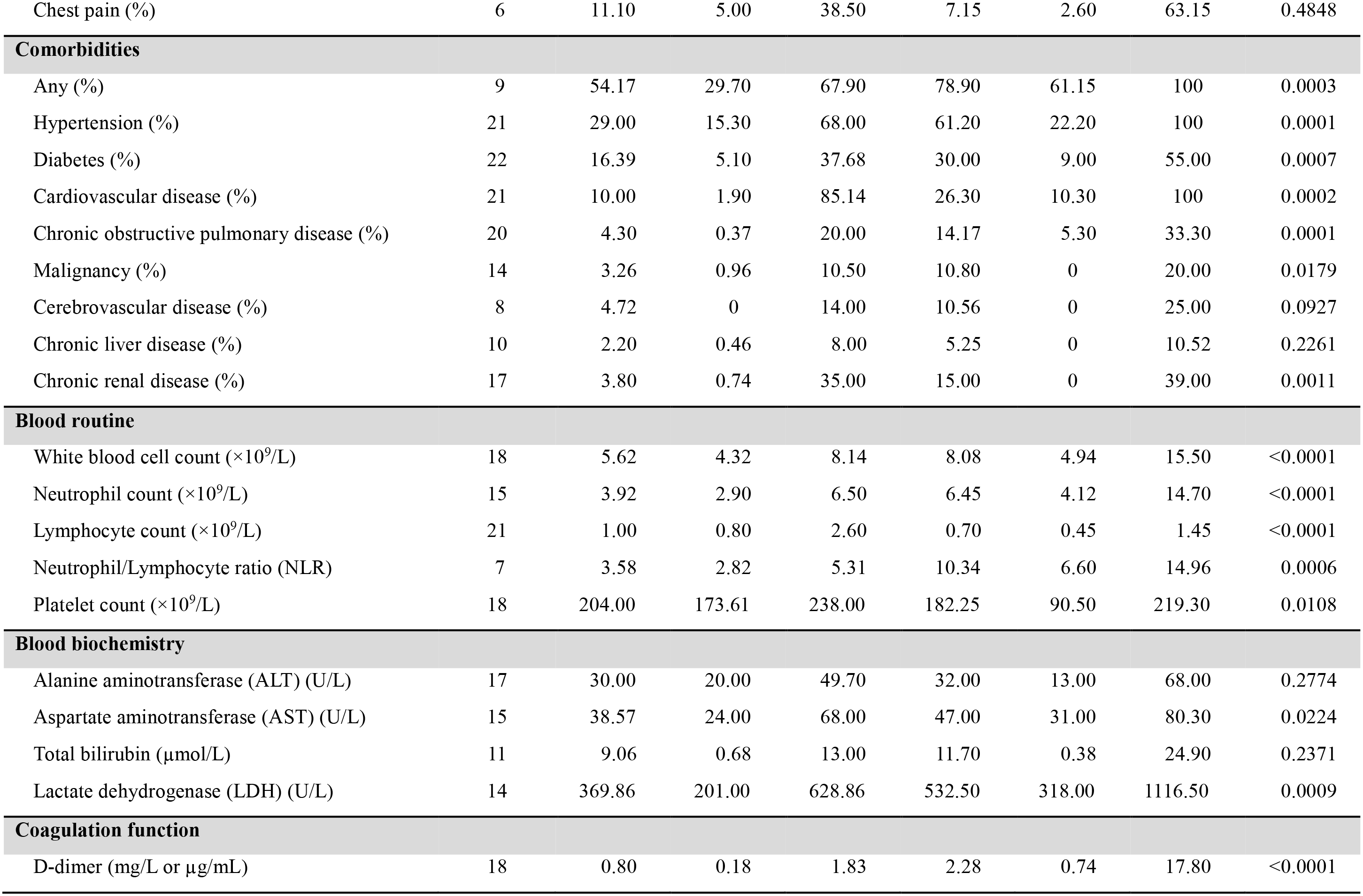
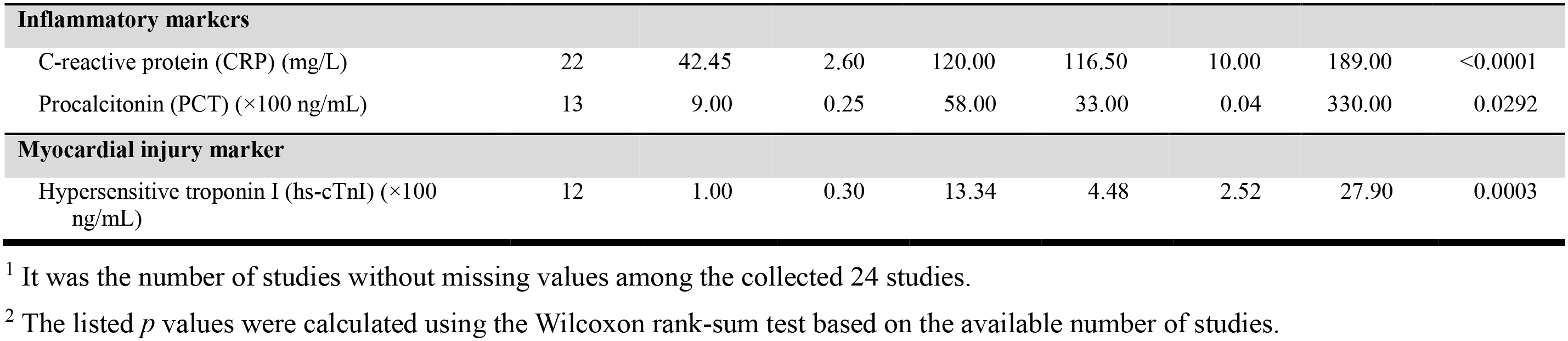
The summary statistics of the demographics, clinical characteristics, and biomarkers of the COVID-19 patients on initial hospital presentations from the collected 24 studies for the assessment of mortality.

